# A Favorable Modifiable Risk Factor Profile Mitigates Polygenic Risk for Alzheimer’s Disease and Related Dementia

**DOI:** 10.64898/2026.06.01.26354634

**Authors:** Clayton O. Mansel, Soniya Mishra, Andrew Craver, Sebastian F. Salathe, John P. Thyfault, Julia A. Bauer, Diego R. Mazzotti, Olivia J. Veatch

## Abstract

**Background:** A recent Lancet Commission estimated that up to 45% of Alzheimer’s Disease and Related Dementias (ADRD) cases could be prevented by addressing modifiable lifestyle risk factors. Meanwhile, genome-wide association studies (GWAS) have shown that common genetic variants also account for substantial ADRD risk. Whether a favorable lifestyle can offset risk in genetically predisposed individuals remains unclear.

**Methods:** We conducted a retrospective cohort study of 105,886 participants from the *All of Us* Research Program enrolled between 2018–2023. Participants were over age 49, assigned male or female at birth, of European ancestry, and without ADRD at baseline. ADRD diagnoses were identified via electronic health records (EHR). Fourteen potentially modifiable risk factors for ADRD were assessed using surveys, EHR records, and wearable data. Genetic risk was quantified as a polygenic risk score (PRS) based on 81 independent GWAS loci and *APOE* ε4 genotype.

**Results:** Overall, 967 incident ADRD events occurred over a median follow-up of 3.7 years. Ten out of 13 modifiable risk factors were significantly associated with ADRD. When grouped into risk factor profiles, intermediate and unfavorable modifiable risk factor scores were associated with substantially higher ADRD risk (HR 3.07, 95% CI 2.47–3.83; HR 8.01, 95% CI 6.39– 10.05, respectively) compared to a favorable lifestyle; APOE ε4 dosage and polygenic risk score were also independently associated with ADRD risk. Among individuals in the highest polygenic risk group, a favorable lifestyle reduced ADRD risk from HR 18.63 (95% CI 10.25–33.86) to 1.90 (95% CI 0.94–3.81), whereas APOE ε4 homozygotes remained at elevated risk even with a favorable lifestyle (HR 6.52, 95% CI 2.97–14.33).

**Conclusions:** Our data suggest ADRD risk is driven more by modifiable risk factors and *APOE* genotype than polygenic risk score. Future genomic-informed risk assessments for ADRD should calibrate their findings to accurately identify high-risk individuals.

## Background

Genomic-informed risk assessments (GIRA) integrate polygenic risk scores, monogenic testing, family history, and traditional clinical risk factors to improve prevention of chronic disease [1– 3]. However, the relationships between genetics and modifiable clinical risk factors within GIRAs remain poorly understood, leading to generic or unclear care recommendations for both patients and healthcare providers [4]. Alzheimer’s Disease and related dementias (ADRD) represent an especially compelling use-case for GIRAs given its high prevalence and economic burden expected to surpass $1.7 trillion dollars globally by 2030 [5]. Moreover, new pharmacological [6,7] and non-pharmacological [8,9] treatments initiated early in the disease course now make identification of high-risk individuals during the decade-long prodromal phase an urgent clinical priority. Large prospective biobanks such as the UK Biobank and *All of Us* Research Program, which link genomic data to lifestyle and environmental variables, offer an unprecedented opportunity to clarify how genetic risk interacts with — or is offset by — modifiable risk factors such as hypertension, diabetes, and depression, ultimately informing more actionable GIRAs for ADRD [10–13]

Previous studies that attempted to examine whether a favorable modifiable risk factor profile mitigates genetic risk for ADRD have reached diverging conclusions [13–15]. It is also unclear if polygenic score screening generalizes to higher-risk patient populations with common co-morbidities in older adults. For example, one previous study concluded that a polygenic score was not associated with cognitive decline specifically in individuals with type II diabetes. To address this need, our study leveraged the *All of Us* Research Program, a prospective U.S.-based, multisite, electronic-health record (EHR)-based dataset with medical records, survey data, and wearable data linked to whole-genome sequencing [10]. We performed a retrospective longitudinal cohort study comparing the relative contribution of both genetic risk (i.e., *APOE* genotype or polygenic risk scores) and modifiable risk factors to risk for incident ADRD. Based on previous studies, we hypothesized that both genetics and modifiable risk factors would provide additive and complementary information to an individual’s risk for ADRD.

## Methods

### All of Us Study Population

The *All of Us* (AoU) Research Program is one of the largest multi-site, longitudinal biorepositories in the world. AoU began enrollment in 2017 and contains survey, electronic health record (EHR), wearable, genomic, and other data [10,16]. EHR data from dozens of healthcare providers across the U.S. are harmonized using the Observational Medical Outcomes Partnership (OMOP) Common Data Model [17]. The data for this study were acquired under the Data Use and Registration Agreement between the University of Kansas Medical Center and *All of Us*. Curated Data Repository v8 (the latest version as of early 2026) was used in the present study. Participants were included if they met the following criteria: 1) were >49 years old at enrollment, 2) reported being assigned male or female at birth, 3) had >1 year available HER data, 4) had available short-read whole-genome sequencing (srWGS) data, and 4) were assigned to European ancestry by AoU. Individuals were excluded if their srWGS data were flagged by AoU, if they had missing data in any of the modifiable risk factor data detailed below, or if they had ADRD at baseline. In total, 105,886 individuals met these criteria.

### Ascertainment of ADRD

ADRD was defined using available EHR records based on a previously validated computable phenotype [18]. This algorithm defined ADRD as the presence of five or more visits on different dates with matching ICD-9/10 codes or 1 matching drug exposure. The exact ICD-9/10 codes or drug exposures are detailed in the Online Methods. Incident ADRD diagnosis date was defined as the date of the 5^th^ visit or drug exposure, whichever occurred earlier.

### Ascertainment of Potentially Modifiable Risk Factors

Potentially modifiable risk factors for ADRD were chosen if they were included in the 2024 *Lancet* Commission on dementia prevention, intervention, and care [19]. We acknowledge debate surrounding the term “modifiable,” as these factors may also be shaped by broader social and structural determinants; however, for conciseness, we refer to them hereafter as “modifiable risk factors.” The *Lancet* Commission identified fourteen modifiable risk factors for dementia including: hearing loss, excessive alcohol use, major depression disorder, traumatic brain injury, type II diabetes, hypertension, low education, low social contact, smoking, obesity, high cholesterol, vision loss, air pollution, and physical activity. Most modifiable risk factors were ascertained using available EHR data except for low education (self-reported), low social contact (self-reported), smoking (self-reported), air pollution (geolocation mapped to historical air pollution data), and physical activity (wearable data). The details of how each modifiable risk factor was ascertained, including which ICD-9/10 codes or survey questions were used is available in the Online Methods. In general, all modifiable risk factors were classified as binary (y/n). The index date for risk factor ascertainment was the date the participant completed the *Overall Health* survey which, in most cases, was the participant’s AoU enrollment date.

### Weighted Modifiable Risk Factor Score

The modifiable risk factor profile of each AoU participant was summarized using a weighted approach. First, an unweighted score was calculated by summing the number of baseline risk factors per participant (Supplementary Figure 1A). Participants were then categorized as favorable (0 risk factors), intermediate (1–2), or unfavorable (≥3). To derive the weighted score, Cox proportional hazards models were fit for each risk factor (see Statistical Analyses), adjusting for age and sex assigned at birth, and the resulting effect sizes were summed. Cutoffs for the weighted score categories were determined by plotting the weighted score against the unweighted score (Supplementary Figure 2). The threshold between intermediate and unfavorable categories was set using the 75th percentile of the unweighted score = 2 group. This approach yielded an approximately normal distribution of weighted modifiable risk factor scores (Supplementary Figure 3).

### Polygenic Risk Score for ADRD

A polygenic risk score (PRS) for ADRD was chosen from the PGS catalogue (catalogue ID #PGS002280) [20,21]. This PRS was chosen because it developed using allele weights from one of the largest to-date genome-wide association study (GWAS) for ADRD comprised of individuals of European ancestry [22]. The PRS contains 83 genome-wide significant loci excluding the *APOE* region. The PRS was calculated in AoU samples using AoU PRS, a cost-effective Python module that computes the PRS directly from the Hail VDS [23]. Raw scores were normalized to z-scores for downstream analyses.

In developing this study, we also attempted to capture polygenic risk in non-European individuals using a multi-ancestry GWAS meta-analysis [24] (PGS003956) and an African ancestry GWAS [25] (PGS004590). However, these polygenic risk scores were not associated with incident ADRD after accounting for population stratification (data not shown). Therefore, we could not compare a PRS to modifiable risk factors in individuals of non-European ancestry.

### Statistical Analyses

Summary statistics are presented as n (%) for categorical variables and mean (standard deviation) for continuous ones. Associations between modifiable risk factors or polygenic risk scores and ADRD were examined using Cox proportional hazards models with time-to-ADRD diagnosis used as the dependent variable. Individuals were censored at last follow-up if the event had not occurred. The proportionality of hazards assumption was assessed using the Schoenfeld residuals technique and was satisfied (p = 0.19). Age, sex assigned at birth, socioeconomic status (SES), and the first ten genomic principal components were included as covariates when appropriate. SES was determined by the AoU-calculated deprivation index score of the AoU participant’s 3-digit zip-code prefix area.

## Results

Overall, the 105,886 AoU participants that met the study inclusion criteria are summarized in Table 1. Participants enrolled in AoU between 05/31/2017 and 09/12/2023. At baseline, the participants had a mean (SD) age of 54.9 (12.4) years and were 43.9% assigned male at birth (Table 1). 75.5% of the participants did not carry an *APOE* ε4 allele. All modifiable risk factors for ADRD were ascertained at baseline using survey data, the most recent lab measures (e.g., BMI), or historical EHR data prior to enrollment (see Online Methods for details). Fourteen modifiable risk factors were chosen because they were included in the 2024 *Lancet* Commission [19]. The prevalence of thirteen out of the fourteen risk factors is summarized in Table 1. A self-reported smoking history was the most prevalent risk factor at 44.0% of the study sample followed closely by high cholesterol (43.7%) and hypertension (43.6%). Physical activity was not included in our study because only 11,485 participants in our cohort had available Fitbit data as (Supplementary Table 1). Over a median (IQR) follow-up of 3.70 (2.95) years, 967 incident ADRD events occurred.

**Table 1.**
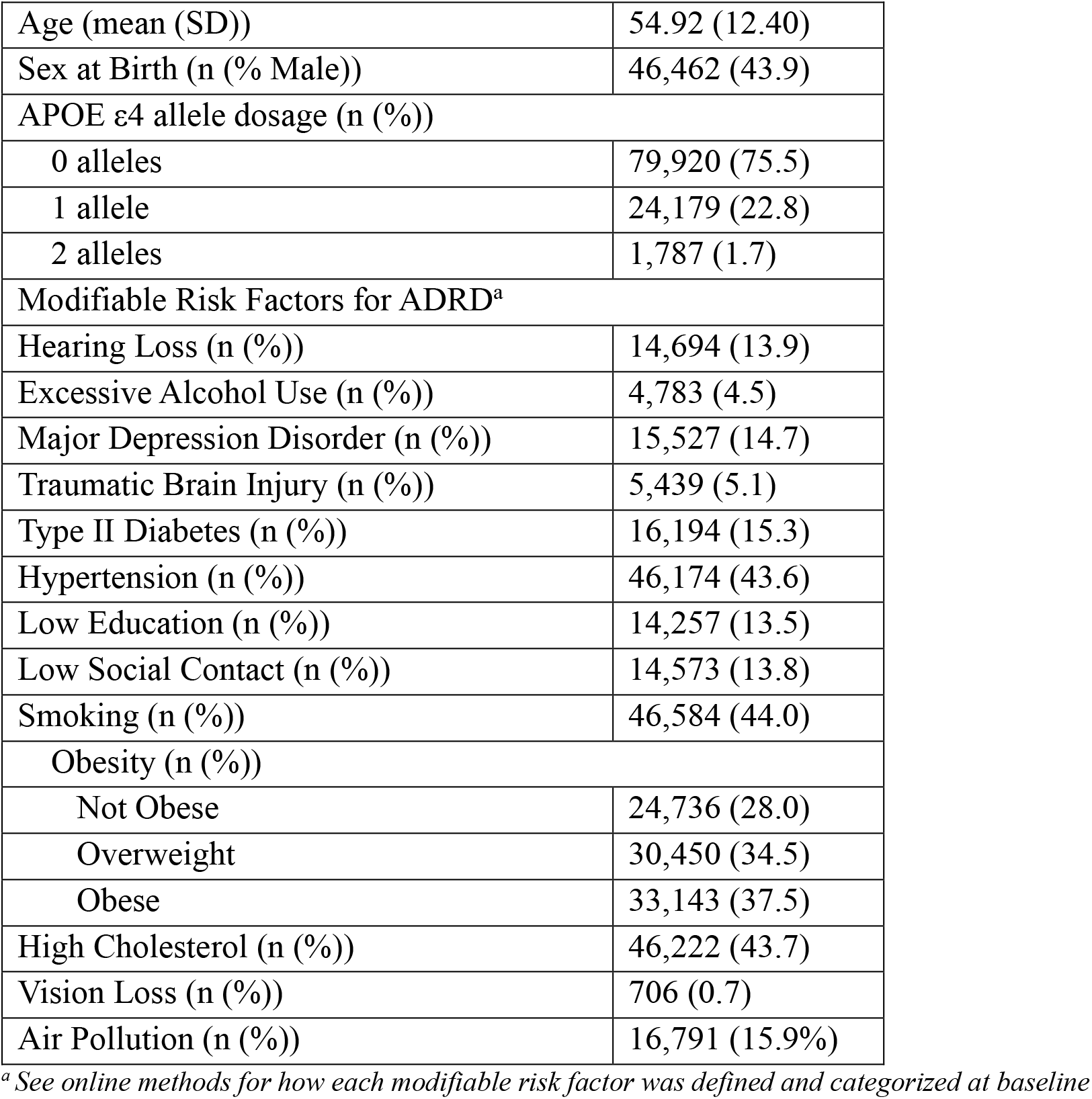
Summary Statistics for 105,886 *All of Us* participants.

|  |  |
| --- | --- |
| Age (mean (SD)) | 54.92 (12.40) |
| Sex at Birth (n (% Male)) | 46,462 (43.9) |
| APOE ε4 allele dosage (n (%)) |  |
| 0 alleles | 79,920 (75.5) |
| 1 allele | 24,179 (22.8) |
| 2 alleles | 1,787 (1.7) |
| Modifiable Risk Factors for ADRD<sup>a</sup> |  |
| Hearing Loss (n (%)) | 14,694 (13.9) |
| Excessive Alcohol Use (n (%)) | 4,783 (4.5) |
| Major Depression Disorder (n (%)) | 15,527 (14.7) |
| Traumatic Brain Injury (n (%)) | 5,439 (5.1) |
| Type II Diabetes (n (%)) | 16,194 (15.3) |
| Hypertension (n (%)) | 46,174 (43.6) |
| Low Education (n (%)) | 14,257 (13.5) |
| Low Social Contact (n (%)) | 14,573 (13.8) |
| Smoking (n (%)) | 46,584 (44.0) |
| Obesity (n (%)) |  |
| Not Obese | 24,736 (28.0) |
| Overweight | 30,450 (34.5) |
| Obese | 33,143 (37.5) |
| High Cholesterol (n (%)) | 46,222 (43.7) |
| Vision Loss (n (%)) | 706 (0.7) |
| Air Pollution (n (%)) | 16,791 (15.9%) |

<sup>a</sup> See online methods for how each modifiable risk factor was defined and categorized at baseline

Out of the thirteen included modifiable risk factors for ADRD, ten were significantly associated with ADRD in the follow-up period (Figure 1; Supplementary Table 2). The most significantly associated risk factor was major depression disorder (HR: 3.48 (95% CI: 3.02-4.00)). Three risk factors were not significantly associated with incident ADRD: smoking, obesity, and air pollution (Figure 1). Further, overweight BMI, obese BMI, and air pollution were protective against incident ADRD with hazard ratios (95% CI) of 0.80 (0.68-0.95), 0.75 (0.63-0.89), and 0.94 (0.91-0.97), respectively. To determine a participant’s modifiable risk factor profile, a weighted modifiable risk score was constructed (see Methods) and this score was categorized into three groups: favorable, intermediate, and unfavorable. Compared to the reference favorable group, both an intermediate (HR: 3.07 (2.47-3.83)) and unfavorable (HR: 8.01 (6.39-10.05)) modifiable risk factor score was significantly associated with ADRD (Figure 2; Supplementary Table 3).

**Figure 1.**
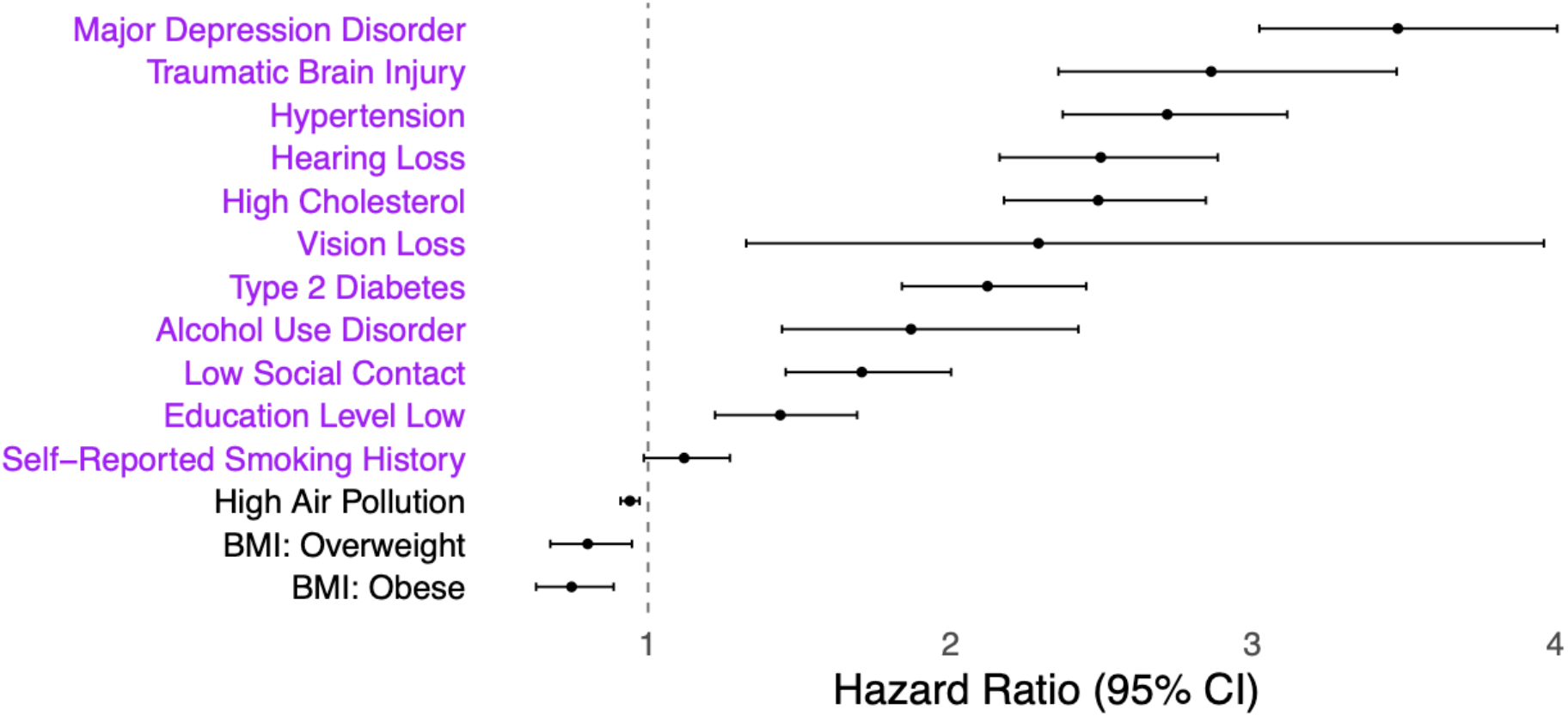
Associations between each potentially modifiable risk factor and incident ADRD. The association between each risk factor and incident ADRD was determined using separate cox proportional hazard models including each binary risk factor and age, sex assigned at birth, and SES (area deprivation index) as covariates. Risk factors shown in purple were later included in a weighted modifiable risk factor score.

**Figure 2.**
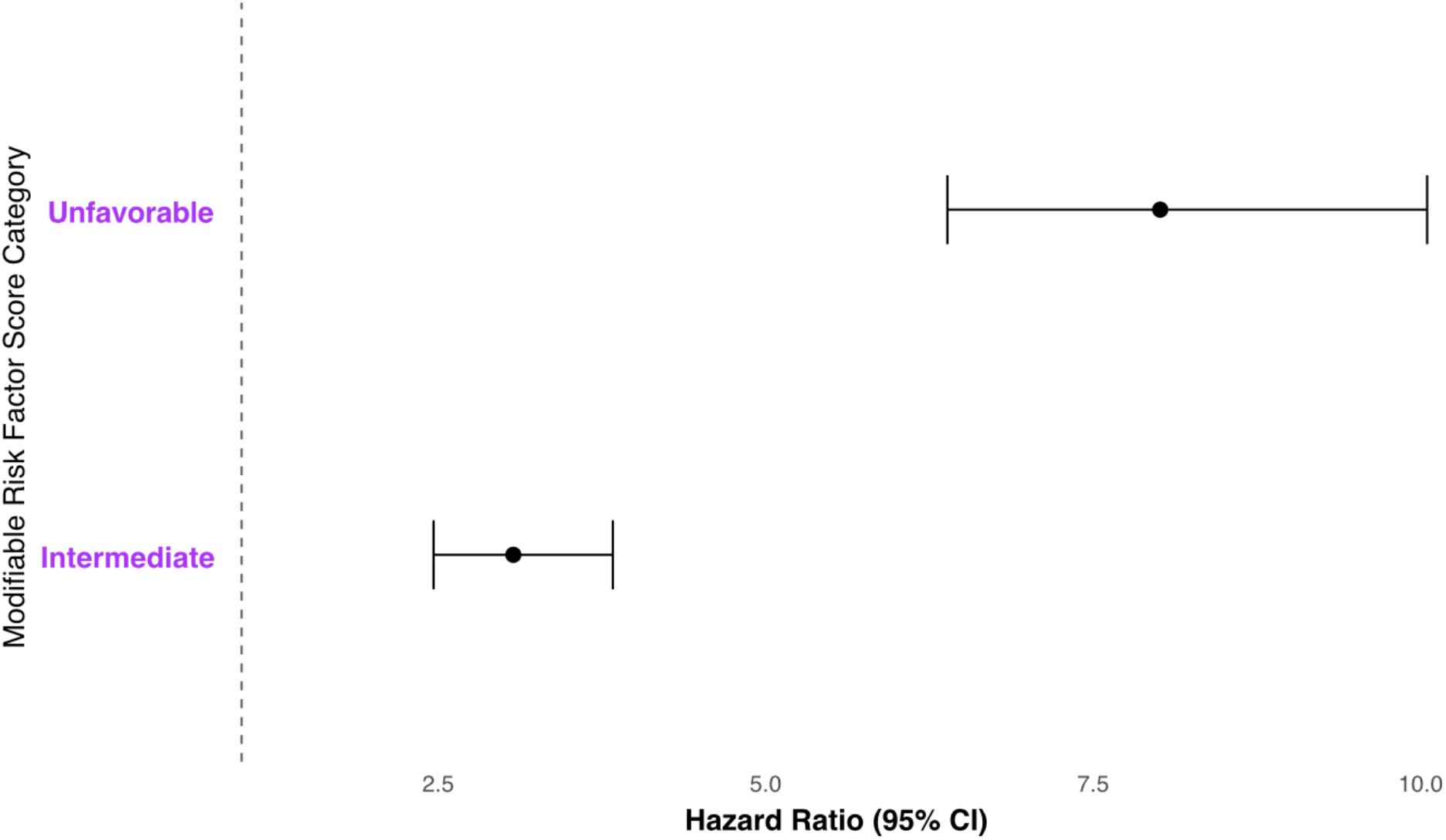
The association between modifiable risk factor score categories and incident ADRD. The association between an individual’s modifiable risk factor profile and incident ADRD was quantified by categorizing modifiable risk factors into a weighted score (see methods). Cox proportional hazard models are adjusted for age, sex assigned at birth, and SES (area deprivation index).

**Table 2.**
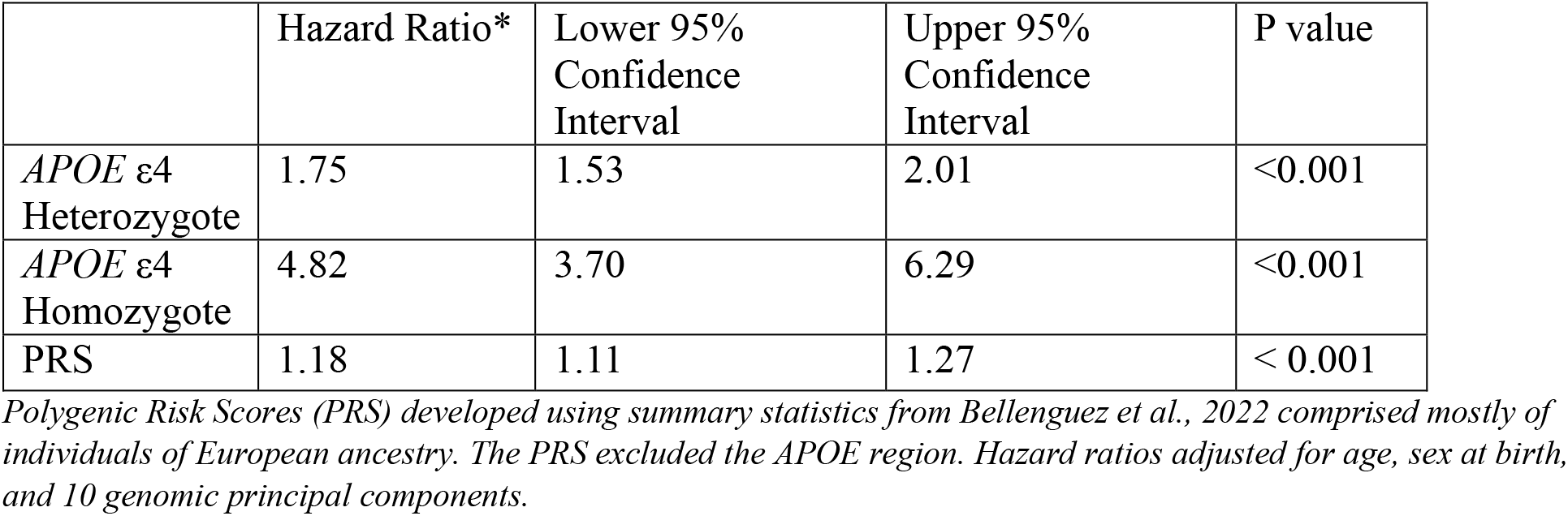
The association between *APOE* genotype and Polygenic Risk Scores for ADRD and incident ADRD.

|  | Hazard Ratio* | Lower 95% Confidence Interval | Upper 95% Confidence Interval | P value |
| --- | --- | --- | --- | --- |
| *APOE* ε4 Heterozygote | 1.75 | 1.53 | 2.01 | <0.001 |
| *APOE* ε4 Homozygote | 4.82 | 3.70 | 6.29 | <0.001 |
| PRS | 1.18 | 1.11 | 1.27 | < 0.001 |

*Polygenic Risk Scores (PRS) developed using summary statistics from Bellenguez et al., 2022 comprised mostly of individuals of European ancestry. The PRS excluded the APOE region. Hazard ratios adjusted for age, sex at birth, and 10 genomic principal components.*

**Table 3.** Interactions between potentially modifiable risk factors and polygenic risk score.

| Risk Factor | p value for interaction term |
| --- | --- |
| Alcohol use disorder | 0.65 |
| MDD | 0.73 |
| T2D | 0.66 |
| Hearing Loss | 0.58 |
| Low Education | 0.69 |
| Low Social Contact | 0.46 |
| High Cholesterol | 0.14 |
| Vision Loss | 0.84 |
| Hypertension | 0.73 |

Interactions between each risk factor determined by adding an interaction term to a cox proportional hazards model including each binarized risk factor, age, sex assigned at birth, and SES (area deprivation index) as covariates. MDD = Major Depressive Disorder, T2D = Type II Diabetes. Significance determined by $p < 0.10$ .

*APOE* genotype was associated with incident ADRD with a hazard ratio of 1.75 (1.53-2.01) and 4.82 (3.70-6.29) for heterozygotes and homozygotes, respectively (Table 2). A one standard deviation increase in PRS was also associated with incident ADRD (HR: 1.18 (1.11-1.27) (Table 2). To examine possible interactions between each modifiable risk factor and a PRS, we included interaction terms in our Cox proportional hazards models, adjusting for age, sex, and SES. Overall, none of the interactions were significant at p < 0.05 (Table 3).

To determine whether a high polygenic risk for ADRD can be mitigated by modifiable risk factors, we stratified our cohort nine ways. First, we categorized the PRS into low (1^st^ quartile), medium (2^nd^ and 3^rd^ quartile), and high (4^th^ quartile). Then, within each PRS category, we stratified by the previously defined weighted risk factor score categories (favorable, intermediate, and unfavorable). Compared to the low PRS/favorable lifestyle reference group, every other strata was significantly associated with ADRD at p <0.05 except for the high PRS/favorable lifestyle strata (Figure 3; Supplementary Table 4). In the highest PRS category, the hazard ratio (95% CI) for ADRD decreased from 18.63 (10.25-33.86) with an unfavorable risk factor score to 1.90 (0.94-3.81) (p=0.07) with a favorable risk factor score.

**Figure 3.**
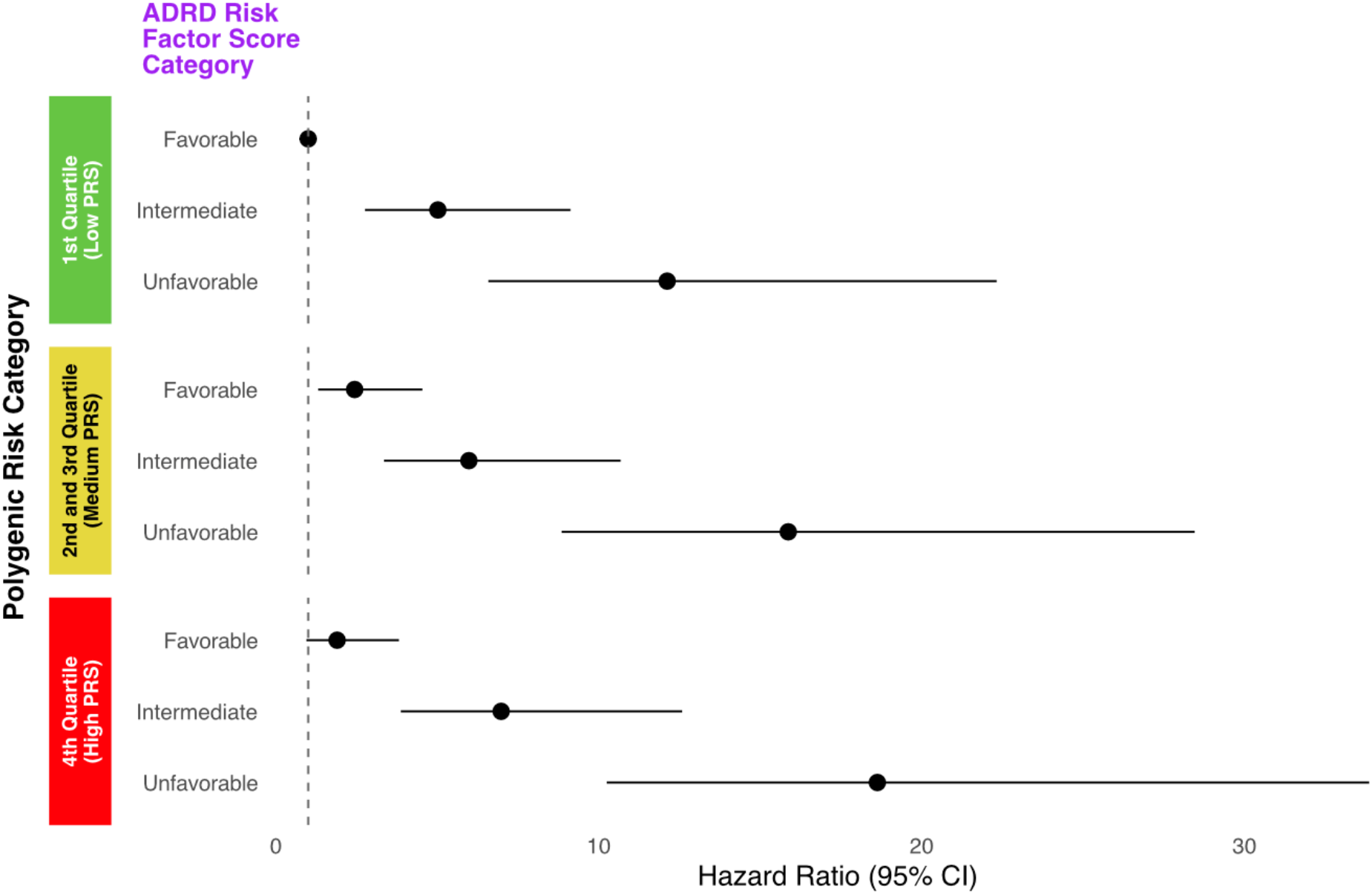
Incident ADRD risk by polygenic and modifiable risk factor category. All of Us participants were stratified by both modifiable risk factor profile (see methods) and polygenic risk score (categorized by quantiles). Using the low PRS/favorable modifiable risk factor group as the reference, cox proportional hazards models quantify the relative risk for ADRD including age, sex assigned at birth, SES (area deprivation index), and 10 genomic principal components as covariates.

To explore whether modifiable risk factors can mitigate risk conferred by *APOE* genotype, we performed a similar analysis but instead stratified by *APOE* ε4 allele dosage (Figure 4; Supplementary Table 5). Compared to the reference group, every other stratum was significantly associated with ADRD at p<0.05. For individuals who inherited 2 *APOE* ε4 alleles, an unfavorable modifiable risk factor profile carried a hazard ratio (95 % CI) of 27.67 (15.67-48.87) while a favorable risk factor profile lowered the hazard ratio (95% CI) to 6.52 (2.97-14.33) (Figure 4; Supplementary Table 5).

**Figure 4.**
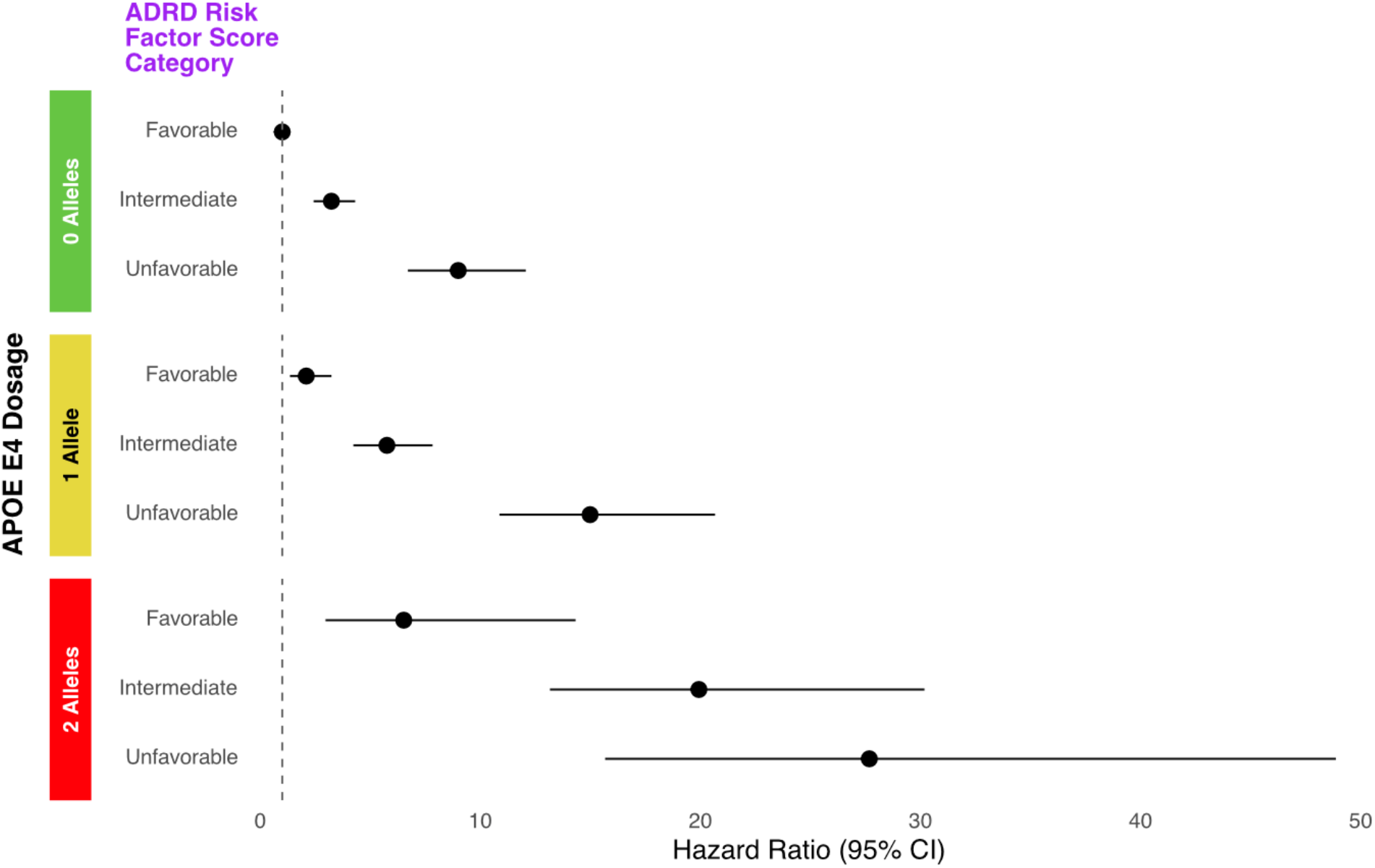
Incident ADRD risk by *APOE* ε4 allele dosage and lifestyle score category. All of Us participants were stratified by both modifiable risk factor profile (see methods) and APOE ε4 genotype (by allele dosage). Using the 0 allele/favorable modifiable risk factor group as the reference, cox proportional hazards models quantify the relative risk for ADRD including age, sex assigned at birth, SES (area deprivation index), and 10 genomic principal components as covariates.

## Discussion

Our data suggest ADRD risk is driven more by modifiable risk factors than polygenic risk. Although high PRS was associated with incident ADRD, this risk was largely mitigated among individuals with favorable lifestyle profiles (HR (95% CI): 1.90 (0.94-3.81)). This pattern was weaker for *APOE* genotype. For example, even with a favorable lifestyle score, individuals in our study with 2 copies of the *APOE* ε4 allele carried a HR (95% CI) of 6.52 (2.97-14.33). Given the high prevalence of undiagnosed prodromal ADRD in primary care [26,27], it is critical that we develop effective strategies for identifying high risk patients. Our data suggest those high-risk patients, at least today, are best identified through the profiling of modifiable risk factors or *APOE* genotype compared to polygenic screening.

During our study, we replicated many ADRD risk factor associations previously identified with effect sizes similar to other studies. For example, one study examined 182,473 dementia-free individuals in the UKBB across 11 modifiable risk factors and found hazard ratios for ADRD that ranged from 1.87 (95% CI: 1.65-2.14) for diabetes to 1.09 (95% CI: 0.89-1.24) for alcohol consumption [28]. Another study (Lourida et al.) that attempted to combine modifiable risk factors into ordinal scores in the UKBB found hazard ratios for ADRD of 1.17 (95% CI: 1.04-1.31) for intermediate and 1.35 (95% CI: 1.15-1.58) for unfavorable healthy lifestyle scores [13]. These estimates are notable lower than our modifiable risk factor score hazard ratio estimates of 3.07 (95% CI; 2.47-3.83) for intermediate and 8.01 (95% CI: 6.39-10.05) for unfavorable. This could be due to a variety of factors including 1) Lourida et al., ascertained only four risk factors (diet, physical activity, alcohol consumption, and smoking) [13], 2) their risk factors were self-reported which could be biased toward a healthier lifestyle, and 3) our estimates may be inflated by summing multiple modifiable risk factors that may interact (e.g., diabetes and hypertension).

Notably, we could not replicate three previous modifiable risk factor associations: physical activity, obesity, and air pollution. Our physical activity ascertainment was limited by the small sample size of AoU participants with available wearable data. We found that both an overweight and obese BMI was protective for ADRD despite previous epidemiological evidence linking obesity and ADRD. The apparent protective association of obesity likely reflects reverse causation due to preclinical weight loss given our relatively short follow-up period [29,30]. We also found a small protective effect of high air pollution on incident ADRD risk. We were limited to the use of 3-digit zip code prefixes which offer a lower degree of granularity than previous epidemiological studies linking air pollution and ADRD often achieving 1km^2 resolutions [31]. Overall, we argue that the risk factors we could not replicate were limited by data availability, follow-up length, and data resolution; their inclusion would have likely not changed our main conclusions.

Multiple previous studies comparing how genetic risk interacts with modifiable risk factors for ADRD and have come to conclusions that differ from ours [13–15,28,32]. Two studies in the UKBB found that a favorable lifestyle or frailty index could slightly mitigate genetic risk for ADRD [13,14]. That is, although a favorable lifestyle reduced ADRD risk among individuals with high genetic risk, their risk remained higher than that of individuals with low genetic risk; in contrast, our findings indicate that a favorable lifestyle reduces risk to a level comparable to the low genetic risk group. Another analysis using the Rotterdam Study data found that as genetic risk increased, the detrimental effects of an unfavorable modifiable risk factor profile were decreased [15]. This was later replicated in the UKBB where another group further showed that the population attributable fraction of modifiable risk factors was decreased in the high ADRD polygenic risk group [28].

How can one explain the diverging conclusions between our study and prior studies? Ours is the first study to use data from the *All of Us* Research Program which could differ from other observational cohorts in terms of both selection bias and exposure heterogeneity. First, while all observational studies are susceptible to selection bias, they differ in degree and direction in ways that likely reshape the joint distribution of genetics, lifestyle, and disease. The *All of Us* dataset recruited participants at healthcare provider organizations and included explicit strategies for an inclusive, targeted recruitment of individuals underrepresented in biomedical research [10]. In contrast, the UKBB invited 9.9 million people by mail and received a response rate of 5.5% and is thus susceptible to previously characterized “healthy-volunteer bias” [33–35]. Consequently, participants in the *All of Us* Research Program have a higher prevalence of chronic disease than both the UKBB and the U.S. general population [36,37]. Selection bias in the UKBB has previously been shown to distort both exposure-disease associations [35,38] and genetic associations [39] due to genetic correlates with UKBB participation. Second, exposure heterogeneity in terms of modifiable risk factors could better highlight the beneficial effects of a favorable lifestyle compared to UKBB and the Rotterdam study. This has been hypothesized for other traits like cardiovascular disease, where a systematic review of multifactorial lifestyle interventions were shown to have stronger beneficial effects in high-risk groups [40]. In the case of UKBB and the Rotterdam study, the effects of a favorable lifestyle may be diluted in a relatively homogenous, health-conscious cohort compared to AoU.

Notably, we compared how a favorable lifestyle could mitigate ADRD risk in terms of both polygenic risk score and *APOE* genotype. We found that while a favorable lifestyle could completely mitigate genetic risk in terms of polygenic risk score, the same was not true for *APOE* genotype. The subgroup with a favorable lifestyle but one or two *APOE* ε4 alleles still carried 2 and 6.5 times increased relative risk for ADRD, respectively. The frequency of *APOE* ε4 heterozygosity and homozygosity varies widely by global genetic ancestry [41,42] but is generally estimated to be 15-20% for ε4 heterozygotes and 2-3% for homozygotes [43] in individuals with European ancestry with higher frequencies in individuals of African ancestry [44]. Thus, we argue that genomic-informed precision medicine for ADRD could achieve benefit by focusing on the effects of even a single gene, as has been recommended by the European Task Force for Brain Health Services [45]. A polygenic risk score explains only a tiny (2%) fraction of ADRD liability in twin studies compared to 11% from *APOE* alone [46]. Focusing on *APOE* alone could mitigate two barriers recently identified in qualitative interviews of providers returning genomic-informed risk assessments (GIRA) to eMERGE participants: 1) GIRA are complex to explain to patients and 2) GIRA come with generic care recommendations [4]. The effect of *APOE* alone appears to be strong enough to both influence comprehensive GIRA containing clinical risk factors and has the potential to influence care decisions. For example, studies have shown that the effect of modifiable risk factors differ by *APOE* genotype [15] and that lifestyle interventions, such as the Mediterranean diet, have profoundly different effects in *APOE* ε4 homozygotes [47]. Overall, our study shows current limitations of polygenic risk scores for GIRA for ADRD; yet it also highlights an opportunity to further study patient-oriented outcomes of widespread *APOE* testing in the general population given its potential to identify high-risk individuals and inform care recommendations.

We note that our study should be interpreted in the context of the following limitations. First, we utilized EHR records to identify ADRD cases based on billing codes and drug exposures which are not as accurate as full clinical evaluations by cognitive neurologists at specialist memory centers. Second, we had to exclude individuals who were not assigned to European genetic ancestry because current polygenic risk scores for ADRD did not transfer to African and American Admixed populations in AoU. This is likely due to differences in allele frequencies across ancestral populations. Future GWAS incorporating more non-European individuals may correct this issue. Third, this was a single observational study in a single population that has not been replicated to date which increases the likelihood of a type I error. Despite these limitations, our study adds to the ongoing debate on the relationship between genetic risk assessments for ADRD and existing modifiable risk factors. Understanding this relationship is critical to accurate risk assessments and the eventual precision prevention of ADRD broadly.

## Acknowledgements/Conflicts/Funding Sources/Consent Statement

### Author contributions

COM, DRM, and OJV were involved in the initial conception and design of the study. SM assisted with phenotyping modifiable risk factors in AoU. COM performed all genomic extractions as well as all statistical analyses. AC and JAB helped clean the air pollution dataset and assisted in the interpretation of the findings. SS and JPT assisted with the wearable data extraction and interpretation of the findings. CM drafted the initial manuscript and received feedback and approval from all authors for the final version.

### Data Availability

All data used in this study can be accessed by qualified researchers at allofus.nih.gov.

## Acknowledgements

We acknowledge *All of Us* participants for their contributions, without whom this research would not have been possible. We also thank the National Institutes of Health’s *All of Us* Research Program for making available the participant and genetic data examined in this study. The All of Us Research Program is supported by the National Institutes of Health, Office of the Director: Regional Medical Centers: 1 OT2 OD026549; 1 OT2 OD026554; 1 OT2 OD026557; 1 OT2 OD026556; 1 OT2 OD026550; 1 OT2 OD 026552; 1 OT2 OD026553; 1 OT2 OD026548; 1OT2 OD026551; 1 OT2 OD026555; IAA #: AOD 16037; Federally Qualified Health Centers: HHSN 263201600085U; Data and Research Center: 5 U2C OD023196; Biobank: 1 U24 OD023121; The Participant Center: U24 OD023176; Participant Technology Systems Center: 1 U24 OD023163; Communications and Engagement: 3 OT2 OD023205; 3 OT2 OD023206; and Community Partners: 1 OT2 OD025277; 3 OT2 OD025315; 1 OT2 OD025337; 1 OT2 OD025276. In addition, the *All of Us* Research Program would not be possible without the partnership of its participants.

