## Supplementary material for "A Favorable Modifiable Risk Factor Profile Mitigates Polygenic Risk for Alzheimer’s Disease and Related Dementia": Online Methods

### Alzheimer's Disease and Related Dementia (ADRD) Definition

ADRD was defined based on the presence of **five or more** visits with matching ICD-9/ICD-10 codes ***or*** 1 drug exposure (see below)

Diagnosis Codes:

| Visits | | | Drug Exposures | |
|---|---|---|---|---|
| Vocab | Code | Name | Drug Name | RxNorm Code |
| ICD-9 | 290.0 | Senile dementia, uncomplicated | Donepezil | 135447 |
| ICD-9 | 290.1 | Presenile dementia, uncomplicated | Aricept | 135446 |
| ICD-9 | 290.11 | Presenile dementia with delirium | Galantamine | 4637 |
| ICD-9 | 290.12 | Presenile dementia with delusional features | Razadyne | 583099 |
| ICD-9 | 290.13 | Presenile dementia with depressive features | Rivastigmine | 183379 |
| ICD-9 | 290.20 | Senile dementia with delusional features, uncomplicated | Exelon | 225807 |
| ICD-9 | 290.21 | Senile dementia with depressive features | Tacrine | 10318 |
| ICD-9 | 290.3 | Senile dementia with delirium | Memantine | 6719 |
| ICD-9 | 290.40 | Vascular dementia, uncomplicated | Namenda | 405206 |
| ICD-9 | 290.41 | Vascular dementia with delirium | | |
| ICD-9 | 290.42 | Vascular dementia with delusions | | |
| ICD-9 | 290.43 | Vascular dementia with depressed mood | | |
| ICD-9 | 291.0 | Alcohol withdrawal delirium | | |
| ICD-9 | 291.1 | Alcohol-induced persisting amnestic disorder | | |
| ICD-9 | 291.2 | Alcohol-induced persisting dementia | | |
| ICD-9 | 292.82 | Drug-induced dementia | | |
| ICD-9 | 294.8 | Other specified persistent mental disorders due to conditions classified elsewhere | | |
| ICD-9 | 294.10 | Dementia in conditions classified elsewhere without behavioral disturbance | | |
| ICD-9 | 294.11 | Dementia in conditions classified elsewhere with behavioral disturbance | | |
| ICD-9 | 331.0 | Alzheimer's disease | | |
| ICD-9 | 331.11 | Pick's disease | | |
| ICD-9 | 331.19 | Other frontotemporal dementia | | |
| ICD-9 | 331.82 | Dementia with Lewy bodies | | |
| ICD-10 | F03.90 | Unspecified dementia without behavioral disturbance | | |
| ICD-10 | F03.91 | Unspecified dementia with behavioral disturbance | | |
| ICD-10 | F01.50 | Vascular dementia without behavioral disturbance | | |
| ICD-10 | F01.51 | Vascular dementia with behavioral disturbance | | |
| ICD-10 | F10.231 | Alcohol dependence with withdrawal delirium | | |
| ICD-10 | F10.96 | Alcohol use, unspecified with alcohol-induced persisting amnestic disorder | | |
| ICD-10 | F10.27 | Alcohol dependence with alcohol-induced persisting dementia | | |
| ICD-10 | F19.97 | Other psychoactive substance use, unspecified with psychoactive substance-induced persisting dementia | | |
| ICD-10 | F06.0 | Psychotic disorder with hallucinations due to known physiological condition | | |

| | | |
|---|---|---|
| ICD-10 | F06.8 | Other specified mental disorders due to known physiological condition |
| ICD-10 | F02.80 | Dementia in other diseases classified elsewhere without behavioral disturbance |
| ICD-10 | F02.81 | Dementia in other diseases classified elsewhere with behavioral disturbance |
| ICD-10 | G30.9 | Alzheimer's disease, unspecified |
| ICD-10 | G31.01 | Pick's disease |
| ICD-10 | G31.83 | Dementia with Lewy bodies |

This algorithm was based on a previously validated computable phenotype with a reported case positive predictive value ranging from 0.73-0.90 and control positive predictive value of 0.97 (Carlson C. Group Health Cooperative. Dementia. PheKB. https://phekb.org/phenotype/10)

### Potentially Modifiable Risk Factors

Potentially modifiable risk factors for ADRD were defined from the 2024 *Lancet* commission on the prevention of dementia [1].

Risk factors were coded as binary (yes/no) based on self-report, physical measurements, or EHR data depending on the risk factor.

The index date for each risk factor was defined as the date the participant completed the *Overall Health* survey which, in most cases, was close to the date that the participant enrolled in *All of Us.*

*All of Us* uses the Observational Medical Outcomes Partnership (OMOP) Common Data Model to store and standardize its EHR data. For each EHR risk factor, the diagnostic codes queried are provided including the OMOP concept ID and ICD9/10 codes. All drug exposures were determined using RXNORM codes.

#### Less Education

Less education was defined as high school or less based on the answer to the following question: What is the highest grade or year of school you completed? (1585940)

#### Hypertension

Hypertension was defined as two or more visits with matching diagnostic codes and 1 or more matching drug exposure (see below)

Diagnostic Codes:

| Name | OMOP Concept ID | Vocab | Code |
|---|---|---|---|
| Benign renovascular hypertension | 44832371 | ICD9CM | 405.11 |
| Benign secondary hypertension | 44831235 | ICD9CM | 405.1 |
| Essential (primary) hypertension | 35207668 | ICD10CM | I10 |
| Essential hypertension | 44833556 | ICD9CM | 401 |
| Hypertension secondary to other renal disorders | 35207676 | ICD10CM | I15.1 |
| Malignant renovascular hypertension | 44831234 | ICD9CM | 405.01 |
| Malignant secondary hypertension | 44831233 | ICD9CM | 405 |
| Other benign secondary hypertension | 44827781 | ICD9CM | 405.19 |

| | | | |
|---|---|---|---|
| Other malignant secondary hypertension | 44824236 | ICD9CM | 405.09 |
| Secondary hypertension | 44832370 | ICD9CM | 405 |
| Secondary hypertension | 1569124 | ICD10CM | I15 |
| Unspecified essential hypertension | 44821949 | ICD9CM | 401.9 |
| Unspecified secondary hypertension | 44837098 | ICD9CM | 405.9 |

Drug Exposure Codes:

| Name | OMOP Concept ID | Vocab | Code |
|---|---|---|---|
| Acebutolol | 1319998 | RXNORM | 149 |
| Aliskiren | 1317967 | RXNORM | 325646 |
| Amlodipine | 1332418 | RXNORM | 17767 |
| Atenolol | 1314002 | RXNORM | 1202 |
| Benazepril | 1335471 | RXNORM | 18867 |
| Betaxolol | 1322081 | RXNORM | 1520 |
| Bisoprolol | 1338005 | RXNORM | 19484 |
| Candesartan | 1351557 | RXNORM | 214354 |
| Captopril | 1340128 | RXNORM | 1998 |
| Carvedilol | 1346823 | RXNORM | 20352 |
| Chlorthalidone | 1395058 | RXNORM | 2409 |
| Clevidipine | 19089969 | RXNORM | 233603 |
| Clonidine | 1398937 | RXNORM | 2599 |
| Diltiazem | 1328165 | RXNORM | 3443 |
| Doxazosin | 1363053 | RXNORM | 49276 |
| Enalapril | 1341927 | RXNORM | 3827 |
| Eplerenone | 1309799 | RXNORM | 298869 |
| Eprosartan | 1346686 | RXNORM | 83515 |
| Felodipine | 1353776 | RXNORM | 4316 |
| Fosinopril | 1363749 | RXNORM | 50166 |
| Guanfacine | 1344965 | RXNORM | 40114 |
| Hydralazine | 1373928 | RXNORM | 5470 |
| Hydrochlorothiazide | 974166 | RXNORM | 5487 |
| Indapamide | 978555 | RXNORM | 5764 |
| Irbesartan | 1347384 | RXNORM | 83818 |
| Isradipine | 1326012 | RXNORM | 33910 |
| Labetalol | 1386957 | RXNORM | 6185 |
| Lisinopril | 1308216 | RXNORM | 29046 |
| Losartan | 1367500 | RXNORM | 52175 |
| Methyldopa | 1305447 | RXNORM | 6876 |
| Metoprolol | 1307046 | RXNORM | 6918 |
| Moexipril | 1310756 | RXNORM | 30131 |
| Nadolol | 1313200 | RXNORM | 7226 |

| | | | |
|---|---|---|---|
| Nebivolol | 1314577 | RXNORM | 31555 |
| Nicardipine | 1318137 | RXNORM | 7396 |
| Nifedipine | 1318853 | RXNORM | 7417 |
| Nisoldipine | 1319880 | RXNORM | 7435 |
| Olmesartan | 40226742 | RXNORM | 321064 |
| Perindopril | 1373225 | RXNORM | 54552 |
| Pindolol | 1345858 | RXNORM | 8332 |
| Prazosin | 1350489 | RXNORM | 8629 |
| Propranolol | 1353766 | RXNORM | 8787 |
| Quinapril | 1331235 | RXNORM | 35208 |
| Ramipril | 1334456 | RXNORM | 35296 |
| Spironolactone | 970250 | RXNORM | 9997 |
| Telmisartan | 1317640 | RXNORM | 73494 |
| Terazosin | 1341238 | RXNORM | 37798 |
| Trandolapril | 1342439 | RXNORM | 38454 |
| Valsartan | 1308842 | RXNORM | 69749 |
| Verapamil | 1307863 | RXNORM | 11170 |

### Traumatic Brain Injury

TBI was defined as the presence of 1 or more visits with matching diagnosis code

| Name | OMOP Concept ID | Vocab | Code |
|---|---|---|---|
| Traumatic brain injury | 4132546 | ICD-10-CM | S06.9X0 |
| Traumatic brain injury with brief loss of consciousness | 4132083 | ICD-10-CM | S06.9X9 |
| Traumatic brain injury with loss of consciousness | 4132082 | ICD-10-CM | S06.9X9 |
| Traumatic brain injury with moderate loss of consciousness | 4133017 | ICD-10-CM | S06.9X9 |
| Traumatic brain injury with no loss of consciousness | 4133715 | ICD-10-CM | S06.9X0 |
| Traumatic brain injury with prolonged loss of consciousness | 4133018 | ICD-10-CM | S06.9X9 |
| Traumatic or nontraumatic brain injury | 4133611 | ICD-10-CM | S06.9X0 |

### Hearing Impairment

Hearing impairment was defined as the presence of 2 or more visits with matching diagnostic codes on different dates:

| Name | OMOP Concept ID | Vocab | Code |
|---|---|---|---|
| Acquired hearing loss | 36715579 | ICD10CM | H91.90 |
| Hearing loss | 377889 | ICD10CM | H91.90 |

### Smoking

Smoking status was determined by the answer to the following yes/no question in the *Lifestyle* survey: Have you smoked at least 100 cigarettes in your entire life? (There are 20 cigarettes in a pack.)? (1585857)

### Low Social Contact

Low social contact was determined using the following survey question: "In general, how would you rate your satisfaction with your social activities and relationships?" from the *Overall Health* survey (1585735) which let participants rate their answer on a 5-point scale: "Excellent", "Very Good", "Good", "Fair", "Poor".

Individuals were determined to have low social contact if they answered "Fair" or "Poor".

### Excessive Alcohol Consumption

Excessive alcohol consumption was determined by the presence of at least 1 diagnostic code for alcohol use disorder:

| Name | OMOP Concept ID | Vocab | Code |
|---|---|---|---|
| Alcohol abuse | 433753 | ICD10CM | F10.10 |
| Alcohol dependence | 435243 | ICD10CM | F10.20 |
| Alcohol intoxication | 4104431 | ICD10CM | F10.129 |
| Alcohol withdrawal syndrome | 375519 | ICD10CM | F10.239 |
| Alcoholism | 4218106 | ICD10CM | F10.20 |

### Depression

Depression was determined by the presence of two or more visits with matching diagnostic codes for major depressive disorder on different dates:

| Name | OMOP Concept ID | Vocab | Code |
|---|---|---|---|
| Major depressive affective disorder, recurrent episode, in full remission | 44827656 | ICD9CM | 296.36 |
| Major depressive affective disorder, recurrent episode, in partial or unspecified remission | 44834589 | ICD9CM | 296.35 |
| Major depressive affective disorder, recurrent episode, mild | 44829923 | ICD9CM | 296.31 |
| Major depressive affective disorder, recurrent episode, moderate | 44831090 | ICD9CM | 296.32 |
| Major depressive affective disorder, recurrent episode, severe, without mention of psychotic behavior | 44835785 | ICD9CM | 296.33 |
| Major depressive affective disorder, recurrent episode, unspecified | 44831089 | ICD9CM | 296.3 |
| Major depressive affective disorder, single episode, in full remission | 44832232 | ICD9CM | 296.26 |
| Major depressive affective disorder, single episode, in partial or unspecified remission | 44822976 | ICD9CM | 296.25 |
| Major depressive affective disorder, single episode, mild | 44824114 | ICD9CM | 296.21 |
| Major depressive affective disorder, single episode, moderate | 44827654 | ICD9CM | 296.22 |
| Major depressive affective disorder, single episode, severe, without mention of psychotic behavior | 44835784 | ICD9CM | 296.23 |
| Major depressive affective disorder, single episode, unspecified | 44822975 | ICD9CM | 296.2 |

| | | | |
|---|---|---|---|
| Major depressive disorder, recurrent severe without psychotic features | 35207158 | ICD10CM | F33.2 |
| Major depressive disorder, recurrent, in partial remission | 45552479 | ICD10CM | F33.41 |
| Major depressive disorder, recurrent, in remission | 1568219 | ICD10CM | F33.4 |
| Major depressive disorder, recurrent, in remission, unspecified | 45547706 | ICD10CM | F33.40 |
| Major depressive disorder, recurrent, mild | 35207156 | ICD10CM | F33.0 |
| Major depressive disorder, recurrent, moderate | 35207157 | ICD10CM | F33.1 |
| Major depressive disorder, recurrent, unspecified | 35207161 | ICD10CM | F33.9 |
| Major depressive disorder, single episode, in full remission | 45547704 | ICD10CM | F32.5 |
| Major depressive disorder, single episode, in partial remission | 45576541 | ICD10CM | F32.4 |
| Major depressive disorder, single episode, mild | 35207150 | ICD10CM | F32.0 |
| Major depressive disorder, single episode, moderate | 35207151 | ICD10CM | F32.1 |
| Major depressive disorder, single episode, severe without psychotic features | 35207152 | ICD10CM | F32.2 |
| Major depressive disorder, single episode, unspecified | 35207155 | ICD10CM | F32.9 |

### Type II Diabetes

The presence of Type II diabetes was determined by the presence of 2 or more visits with matching diagnostic codes **and** 1 or more matching drug exposure:

Type II Diabetes Diagnosis Codes:

| Name | OMOP Concept ID | Vocab | Code |
|---|---|---|---|
| Diabetes mellitus without mention of complication, type II or unspecified type, not stated as uncontrolled | 44836914 | ICD9CM | 250 |
| Diabetes mellitus without mention of complication, type II or unspecified type, uncontrolled | 44836915 | ICD9CM | 250.02 |
| Diabetes with hyperosmolarity, type II or unspecified type, not stated as uncontrolled | 44836916 | ICD9CM | 250.2 |
| Diabetes with hyperosmolarity, type II or unspecified type, uncontrolled | 44824073 | ICD9CM | 250.22 |
| Diabetes with neurological manifestations, type II or unspecified type, not stated as uncontrolled | 44828795 | ICD9CM | 250.6 |
| Diabetes with neurological manifestations, type II or unspecified type, uncontrolled | 44833366 | ICD9CM | 250.62 |
| Diabetes with ophthalmic manifestations, type II or unspecified type, not stated as uncontrolled | 44819500 | ICD9CM | 250.5 |
| Diabetes with ophthalmic manifestations, type II or unspecified type, uncontrolled | 44829879 | ICD9CM | 250.52 |
| Diabetes with other coma, type II or unspecified type, not stated as uncontrolled | 44826460 | ICD9CM | 250.3 |
| Diabetes with other coma, type II or unspecified type, uncontrolled | 44832193 | ICD9CM | 250.32 |
| Diabetes with other specified manifestations, type II or unspecified type, not stated as uncontrolled | 44831047 | ICD9CM | 250.8 |
| Diabetes with other specified manifestations, type II or unspecified type, uncontrolled | 44826461 | ICD9CM | 250.82 |
| Diabetes with peripheral circulatory disorders, type II or unspecified type, not stated as uncontrolled | 44827616 | ICD9CM | 250.7 |

| | | | |
|---|---|---|---|
| Diabetes with peripheral circulatory disorders, type II or unspecified type, uncontrolled | 44833367 | ICD9CM | 250.72 |
| Diabetes with renal manifestations, type II or unspecified type, not stated as uncontrolled | 44831045 | ICD9CM | 250.4 |
| Diabetes with renal manifestations, type II or unspecified type, uncontrolled | 44832194 | ICD9CM | 250.42 |
| Diabetes with unspecified complication, type II or unspecified type, not stated as uncontrolled | 44827617 | ICD9CM | 250.9 |
| Diabetes with unspecified complication, type II or unspecified type, uncontrolled | 44829882 | ICD9CM | 250.92 |
| Type 2 diabetes mellitus with diabetic cataract | 45595798 | ICD10CM | E11.36 |
| Type 2 diabetes mellitus with diabetic dermatitis | 45547626 | ICD10CM | E11.620 |
| Type 2 diabetes mellitus with diabetic nephropathy | 45591027 | ICD10CM | E11.21 |
| Type 2 diabetes mellitus with diabetic neuropathy, unspecified | 45605403 | ICD10CM | E11.40 |
| Type 2 diabetes mellitus with diabetic peripheral angiopathy without gangrene | 45533021 | ICD10CM | E11.51 |
| Type 2 diabetes mellitus with foot ulcer | 45581355 | ICD10CM | E11.621 |
| Type 2 diabetes mellitus with hyperglycemia | 45605405 | ICD10CM | E11.65 |
| Type 2 diabetes mellitus with hyperosmolarity with coma | 45586139 | ICD10CM | E11.01 |
| Type 2 diabetes mellitus with hyperosmolarity without nonketotic hyperglycemic-hyperosmolar coma (NKHHC) | 45542738 | ICD10CM | E11.00 |
| Type 2 diabetes mellitus with hypoglycemia with coma | 45561949 | ICD10CM | E11.641 |
| Type 2 diabetes mellitus with hypoglycemia without coma | 45591031 | ICD10CM | E11.649 |
| Type 2 diabetes mellitus with other diabetic arthropathy | 45586140 | ICD10CM | E11.618 |
| Type 2 diabetes mellitus with other diabetic kidney complication | 45605401 | ICD10CM | E11.29 |
| Type 2 diabetes mellitus with other diabetic ophthalmic complication | 45533019 | ICD10CM | E11.39 |
| Type 2 diabetes mellitus with other oral complications | 45566731 | ICD10CM | E11.638 |
| Type 2 diabetes mellitus with other skin complications | 45547627 | ICD10CM | E11.628 |
| Type 2 diabetes mellitus with other skin ulcer | 45600642 | ICD10CM | E11.622 |
| Type 2 diabetes mellitus with other specified complication | 45595799 | ICD10CM | E11.69 |
| Type 2 diabetes mellitus with periodontal disease | 45533023 | ICD10CM | E11.630 |
| Type 2 diabetes mellitus with unspecified complications | 35206881 | ICD10CM | E11.8 |
| Type 2 diabetes mellitus with unspecified diabetic retinopathy with macular edema | 45581352 | ICD10CM | E11.311 |
| Type 2 diabetes mellitus with unspecified diabetic retinopathy without macular edema | 45581353 | ICD10CM | E11.319 |
| Type 2 diabetes mellitus without complications | 35206882 | ICD10CM | E11.9 |

Type II Diabetes Drugs:

| Name | OMOP Concept ID | Vocab | Code |
|---|---|---|---|
| Acarbose | 1529331 | RXNORM | 16681 |
| Albiglutide | 44816332 | RXNORM | 1534763 |
| Alogliptin | 43013884 | RXNORM | 1368001 |
| Canagliflozin | 43526465 | RXNORM | 1373458 |
| Chlorpropamide | 1594973 | RXNORM | 2404 |
| Colesevelam | 1518148 | RXNORM | 141626 |

| | | | |
|---|---|---|---|
| Dapagliflozin | 44785829 | RXNORM | 1488564 |
| Dulaglutide | 45774435 | RXNORM | 1551291 |
| Empagliflozin | 45774751 | RXNORM | 1545653 |
| Exenatide | 1583722 | RXNORM | 60548 |
| Glimepiride | 1597756 | RXNORM | 25789 |
| Glipizide | 1560171 | RXNORM | 4821 |
| Glyburide | 1559684 | RXNORM | 4815 |
| Linagliptin | 40239216 | RXNORM | 1100699 |
| Liraglutide | 40170911 | RXNORM | 475968 |
| Lixisenatide | 44506754 | RXNORM | 1440051 |
| Metformin | 1503297 | RXNORM | 6809 |
| Miglitol | 1510202 | RXNORM | 30009 |
| Nateglinide | 1502826 | RXNORM | 274332 |
| Pioglitazone | 1525215 | RXNORM | 33738 |
| Repaglinide | 1516766 | RXNORM | 73044 |
| Rosiglitazone | 1547504 | RXNORM | 84108 |
| Saxagliptin | 40166035 | RXNORM | 857974 |
| Semaglutide | 793143 | RXNORM | 1991302 |
| Sitagliptin | 1580747 | RXNORM | 593411 |
| Tolazamide | 1502809 | RXNORM | 10633 |
| Troglitazone | 1515249 | RXNORM | 72610 |

### Obesity

Obesity was defined using BMI calculated from the height and weight measurements. Height and weight records were obtained at least within 1 year of the index date. A BMI >= 25 but <30 was categorized as overweight and a BMI >30 was categorized as obese.

### High Cholesterol

High cholesterol was defined as two or more visits with matching diagnostic codes on different dates:

| Name | OMOP Concept ID | Vocab | Code |
|---|---|---|---|
| Hyperlipidemia | 432867 | ICD10CM | E78.5 |
| Mixed hyperlipidemia | 438720 | ICD10CM | E78.2 |

### Vision Loss

Vision loss was defined as two or more visits with matching diagnostic codes on different dates:

| Name | OMOP Concept ID | Vocab | Code |
|---|---|---|---|
| Blindness AND/OR vision impairment level | 4023310 | ICD10CM | H54.7 |

### Physical Inactivity

Physical activity was ascertained using linked Fitbit data (step count per day) when available. Step counts less than 200 or greater than 50,000 per day were excluded as implausible. Individuals with less than 30 days of step count data were excluded. Low physical activity was assigned to a participant if they averaged fewer than 5,000 steps per day.

#### Air Pollution

Air pollution exposures were defined using annual surface $PM_{2.5}$ levels estimated using a satellite-derived model from the Atmospheric Composition Analysis Group at Washington University in St. Louis [2–4]. We averaged the $PM_{2.5}$ levels from 2016-2022 and mapped them to the 3-digit zipcode that AoU provides for each consenting participant. High air pollution was defined as a one standard deviation increase in $PM_{2.5}$ levels.
