## Supplementary Material for "A Favorable Modifiable Risk Factor Profile Mitigates Polygenic Risk for Alzheimer’s Disease and Related Dementia"

**Supplementary Table 1. Summary of available Fitbit wearable data stratified by ADRD status**

| | No ADRD (n=11,453) | ADRD (n=32) |
|---|---|---|
| Average Steps per Day (mean (SD)) | 7,058 (3,513) | 5,694 (3,130) |
| Number of Fitbit Days (mean (SD)) | 823 (950) | 821 (831) |
| Categorized as Low Physical Activity (n (%)) | 8,009 (69.9) | 16 (50.0) |

**Supplemental Table 2. The association between lifestyle factors and incident ADRD**

| Risk Factor | Hazard Ratio (95% CI) | p value |
|---|---|---|
| Obese BMI (>30) | 0.75 (0.63-0.89) | <0.001 |
| Overweight BMI (>25<30) | 0.80 (0.68-0.95) | 0.009 |
| Air Pollution (1 SD increase in $PM_{25}$ levels) | 0.94 (0.91-0.97) | <0.001 |
| Smoking | 1.12 (0.99-1.27) | 0.082 |
| Low education | 1.44 (1.22-1.69) | <0.001 |
| Low Social Contact | 1.71 (1.46-2.00) | <0.001 |
| Alcohol Use Disorder | 1.87 (1.44-2.42) | <0.001 |
| Type II Diabetes | 2.12 (1.84-2.45) | <0.001 |
| Vision Loss | 2.29 (1.32-3.96) | 0.003 |
| High Cholesterol | 2.49 (2.18-2.84) | <0.001 |
| Hearing Loss | 2.50 (2.16-2.88) | <0.001 |
| Hypertension | 2.72 (2.37-3.11) | <0.001 |
| Traumatic Brain Injury | 2.86 (2.36-3.48) | <0.001 |
| Major Depression Disorder | 3.48 (3.02-4.00) | <0.001 |

The association between each risk factor and incident ADRD was modelled via independent Cox proportional hazards models adjusted for age and sex.

**Supplemental Table 3. The association between weighted lifestyle score category and incident ADRD**

| Category (ref = Favorable) | Hazard Ratio (95% CI) | p value |
|---|---|---|
| Intermediate | 3.07 (2.47-3.83) | <0.001 |
| Unfavorable | 8.01 (6.39-10.05) | <0.001 |

**Supplemental Table 4. Incident ADRD risk stratified by PRS and weighted lifestyle risk score categories**

| PRS Group | Lifestyle Group | Hazard Ratio (95% CI) | p value |
|---|---|---|---|
| 1st Quartile (Low PRS) | Favorable | ref | ref |
| 2nd and 3rd Quartile (Medium PRS) | Favorable | 2.43 (1.31-4.53) | 0.005 |
| 4th Quartile (High PRS) | Favorable | 1.90 (0.94-3.81) | 0.073 |
| 1st Quartile (Low PRS) | Intermediate | 5.02 (2.76-9.13) | <0.001 |
| 2nd and 3rd Quartile (Medium PRS) | Intermediate | 5.98 (3.35-10.68) | <0.001 |
| 4th Quartile (High PRS) | Intermediate | 6.98 (3.87-12.58) | <0.001 |
| 1st Quartile (Low PRS) | Unfavorable | 12.12 (6.57-22.32) | <0.001 |
| 2nd and 3rd Quartile (Medium PRS) | Unfavorable | 15.87 (8.85-28.46) | <0.001 |
| 4th Quartile (High PRS) | Unfavorable | 18.63 (10.25-33.86) | <0.001 |

**Supplemental Table 5. Incident ADRD risk stratified by *APOE* ε4 allele dosage and weighted lifestyle risk score categories**

| *APOE* ε4 allele dosage | Lifestyle Group | Hazard Ratio (95% CI) | p.value |
|---|---|---|---|
| 0 Alleles | Favorable | ref | ref |
| 1 Allele | Favorable | 2.09 (1.35-3.24) | <0.001 |
| 2 Alleles | Favorable | 6.52 (2.97-14.33) | <0.001 |
| 0 Alleles | Intermediate | 3.23 (2.42-4.31) | <0.001 |
| 1 Allele | Intermediate | 5.76 (4.23-7.83) | <0.001 |
| 2 Alleles | Intermediate | 19.93 (13.16-30.18) | <0.001 |
| 0 Alleles | Unfavorable | 9.00 (6.71-12.07) | <0.001 |
| 1 Allele | Unfavorable | 14.99 (10.87-20.67) | <0.001 |
| 2 Alleles | Unfavorable | 27.67 (15.67-48.87) | <0.001 |

**Supplementary Figure 1: Distribution of unweighted lifestyle score and score categories.**

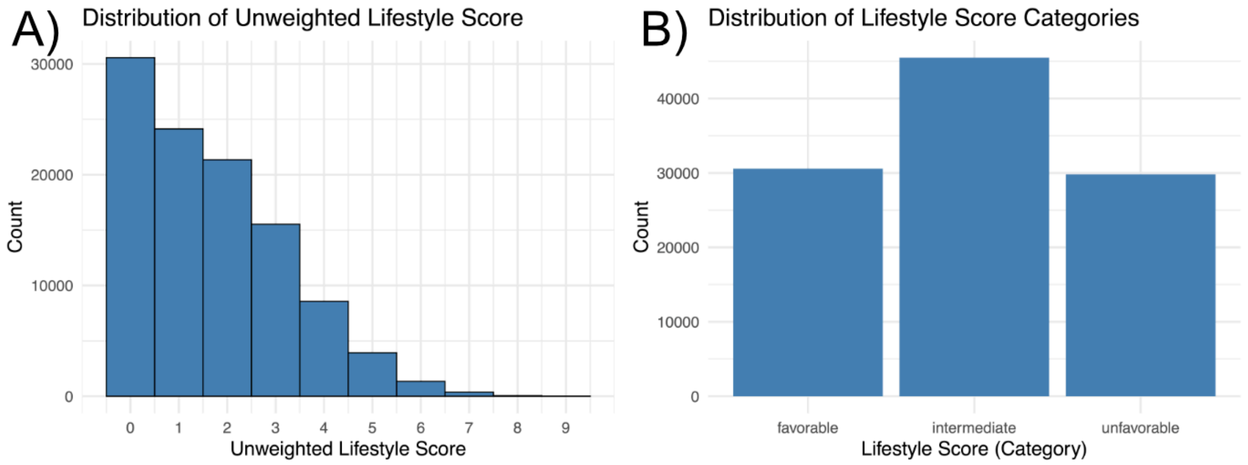

**Supplementary Figure 2: Distribution of weighted lifestyle score by unweighted score.**

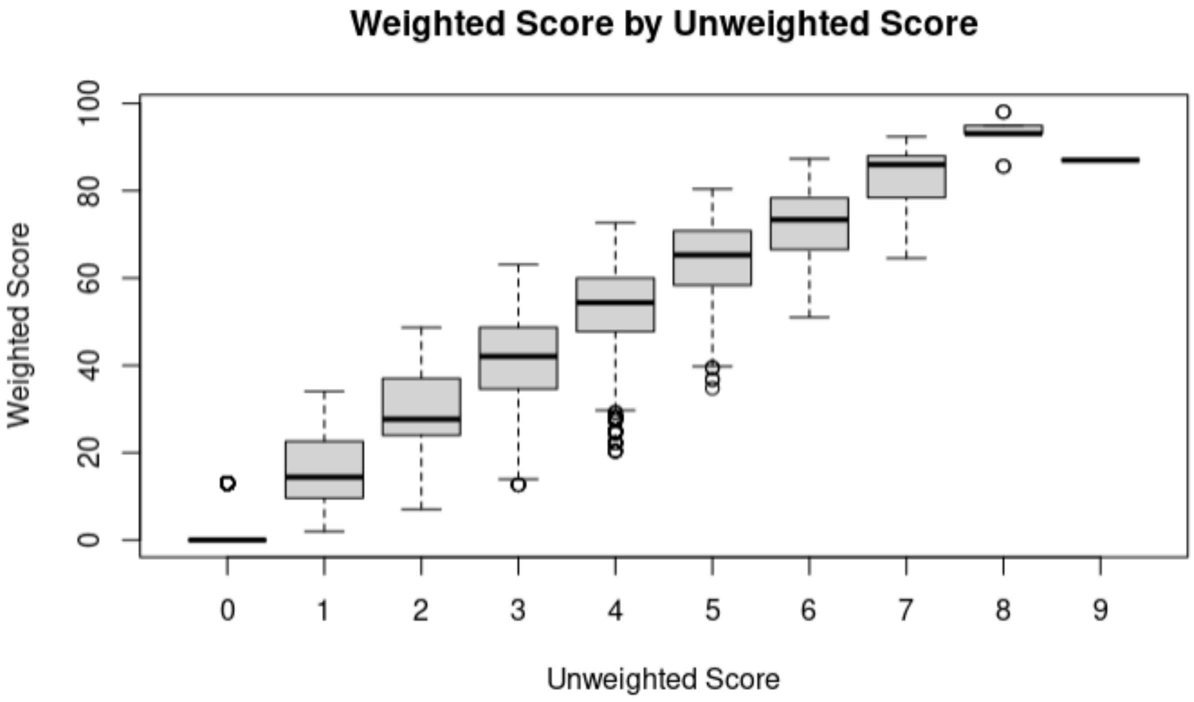

**Supplementary Figure 3: Distribution of weighted lifestyle score categories**

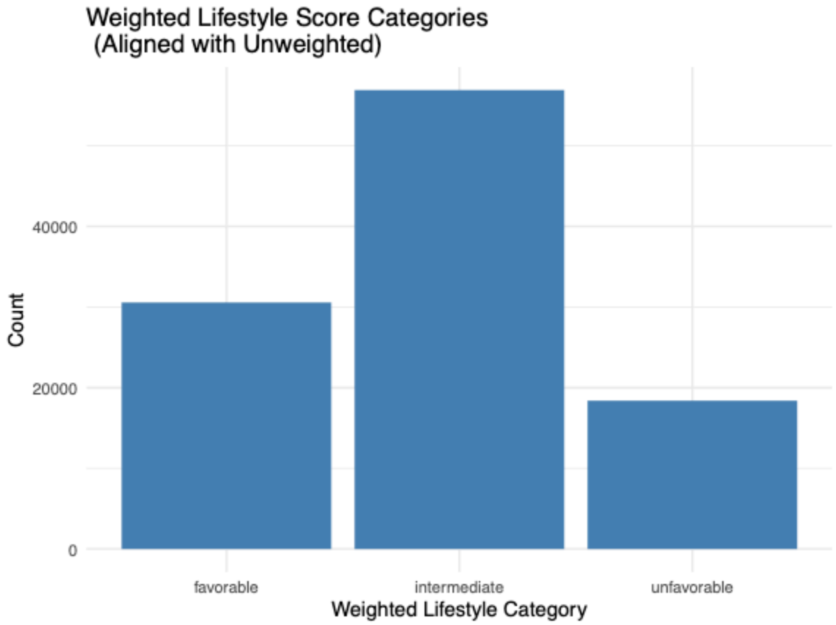
